# Standardized Measurement of Type 1 Diabetes Polygenic Risk Across Multi-Ancestry Population Cohorts

**DOI:** 10.1101/2025.01.16.25320691

**Authors:** Amber M. Luckett, Richard A. Oram, Aaron J. Deutsch, Hector I Ortega, Diane P. Fraser, Kaavya Ashok, Alisa K. Manning, Josep M. Mercader, Manuel Rivas, Miriam S. Udler, Michael N Weedon, Anna L. Gloyn, Seth A. Sharp

**Author notes:** **Corresponding author:** Seth A. Sharp.

## Abstract

Type 1 diabetes (T1D) polygenic risk scores (PRS) are effective tools for discriminating T1D from other diabetes types and predicting T1D risk, with applications in screening and intervention trials. A previously published T1D Genetic Risk Score 2 (GRS2) is widely adopted, but challenges in standardization and accessibility have hindered broader clinical and research utility. To address this, we introduce GRS2x, a standardized and cross-compatible method for accurate T1D PRS calculation, demonstrating genotyping and reference panel independent performance across diverse datasets. GRS2x as a unified approach facilitates accessible and portable measurement of T1D polygenic risk.

## Body

Type 1 diabetes (T1D) polygenic risk scores (PRS) effectively discriminate T1D from both Type 2 (T2D) and monogenic diabetes^1^ and can aid prediction of T1D, serving as a valuable tool in ongoing screening and intervention trials^2^. The T1D Genetic Risk Score 2 (GRS2)^3^ has been most widely adopted, yet absence of a standardized and accessible method of generation has proved a barrier to broader clinical and research utility. Study-specific factors, such as genotyping technology (whole genome sequencing [WGS] vs array), and access to imputation panels, limit the availability of key single nucleotide variants (SNV), particularly in the human leukocyte antigen (HLA) region. Consequently, comparisons across diverse cohorts are constrained, prohibiting standardized interpretation. To address this, we present GRS2x, a cross-compatible and standardized approach for accurate calculation of T1D polygenic risk.

We selected robust SNVs for GRS2x (replacing 18 out of 67 from GRS2) from post-quality control reference panels (1000 Genomes [1000G], Haplotype Reference Consortium [HRC], Trans-Omics for Precision Medicine [TOPMED] r2/3) in linkage disequilibrium with HLA alleles as previously described^3^. Insertions and deletions (>2 base pairs), multinucleated, and structural variants were excluded. SNVs were partitioned by locus (HLA, 6p21.3) and proximal gene (HLA Class II, HLA Class I, and non-HLA) defining partitioned polygenic scores (pPS), with total GRS2x calculated as their sum (Fig. 1A). We addressed critical limitations through the introduction of two algorithms: rank-based removal of excess HLA DR-DQ haplotypes (>2) resulting from SNV introduced noise, and imputation of missing variant estimated contribution, applying Hardy-Weinberg equilibrium with TOPMED reference frequency. Enabled by these approaches, dataset-independent fixed-value normalization was applied with minimum and maximum GRS2x relative-risk values.

**Figure 1:**
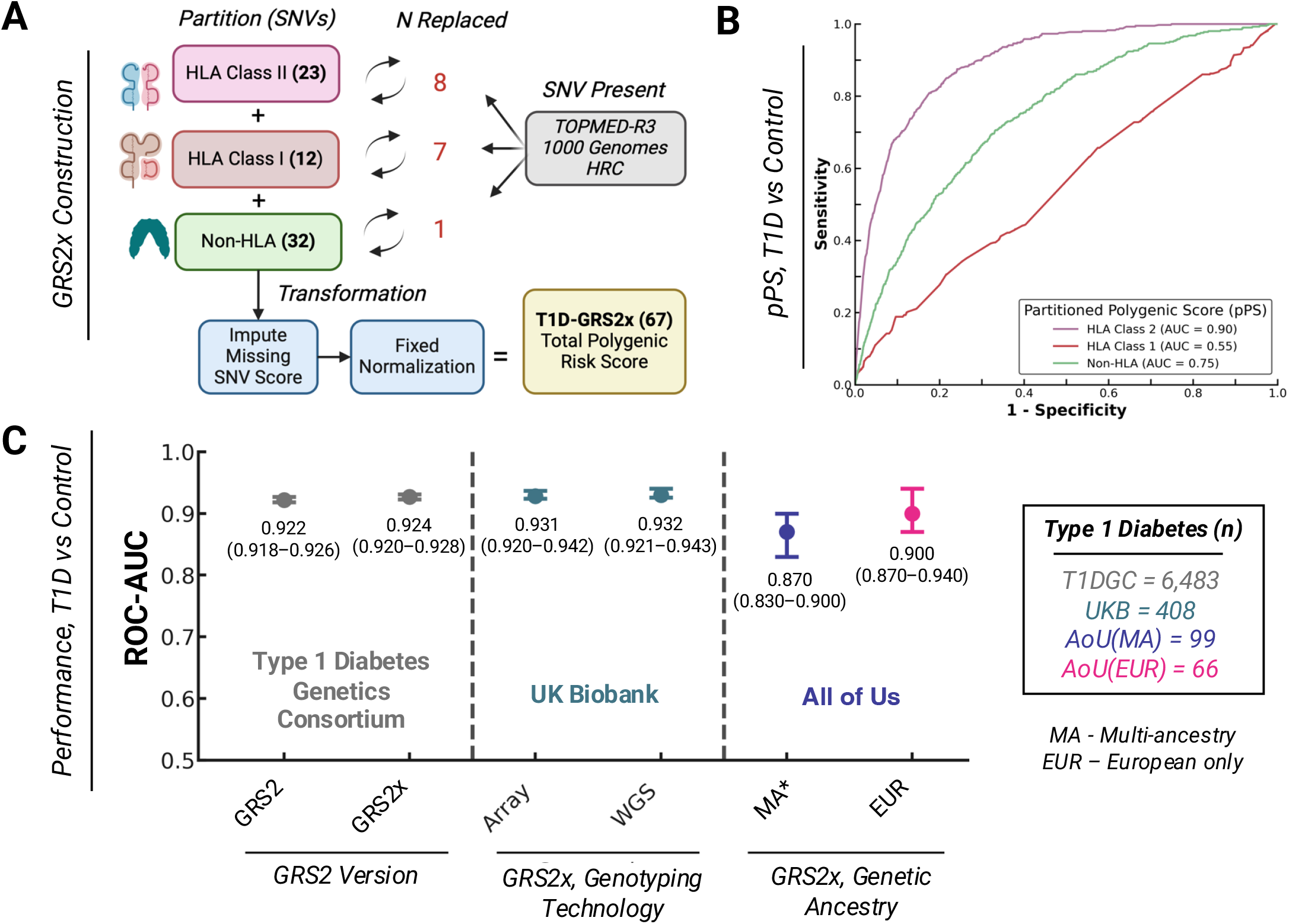
**(A)** Construction of the GRS2x is the sum of partitions defined by locus, after application of missing SNV score imputation and normalization, **(B)** Partitioned Polygenic Scores (pPS) are independently discriminative of T1D in European ancestry UK Biobank individuals with whole-genome sequencing data, **(C)** GRS2x discriminative performance is comparable to GRS2, but independent of genotyping technology (UK Biobank), and consistent across multi-ancestry datasets (All of Us).

Discriminative performance of GRS2x was imputation reference panel independent in array data (ImmunoChip) from The Type 1 Diabetes Genetics Consortium (T1DGC) study. Discrimination of T1D cases from controls (T1D=6,483, Control=9,247; previously defined^3^) imputed with TOPMED(r2) improved marginally over GRS2 (AUROC, GRS2x=0.924 vs GRS2=0.922, p=2.96×10^−10^), equivalent to previously reported performance of GRS2 imputed with 1000G+HRC reference panels (Fig 1C)^3^.

To demonstrate GRS2x performance was independent of genotyping technology and SNV availability, we replicated published T1D and control definitions^3^ in electronic health records (EHR) on the UK Biobank (UKB) Research Analysis Platform (RAP), including European ancestry individuals with genomic data (T1D=408, Control=365,622)^4^. A T1D definition (T1D in EHR, age at first EHR record<30, insulin prescribed, not prescribed non-insulin diabetes medications) was generated on the All of Us (AoU) RAP (T1D=99, Control= 205,943)^5^. Age at EHR record was used as a proxy for juvenile T1D onset due to the absence of pediatric records. Whilst highly specific, T1D definitions are less sensitive due to documented limitations of EHR data. GRS2x performance was measured by Area Under the Receiver Operating Characteristic Curve (AUROC).

T1D case-control discrimination for GRS2x in European-ancestry individuals in UKB (n=367,318, Axiom, HRC imputed) and/or WGS (n=366,030) data was not significantly altered (AUROC, array=0.931 vs WGS=0.932, p=0.9) despite non-overlapping SNVs (n=3) (Fig. 1C). In individuals with both array and WGS genotypes, GRS2x was highly concordant (R=0.96), and individual risk estimates by rank were closely matched (ρ=0.96). Excess HLA DR-DQ genotypes (>2) were resolved in previously unscored individuals (1.14%, n=5550) by the rank-based algorithm. Performance of pPS suggests the majority of T1D case/control discriminative power is attributed to HLA Class II (AUROC=0.90) as compared to HLA Class I or non-HLA risk (AUROC=0.55, 0.75 respectively) (Fig. 1B)

Overall multi-ancestry (MA) T1D case vs control discrimination (AoU) was marginally reduced compared to European ancestry (AUROC, MA=0.870, EUR=0.904, p=0.21) though not statistically different with EHR limited case numbers (Fig. 1C). Furthermore, rank-based concordance of GRS2x vs GRS2 was marginally lower in the MA cohort (ρ_EUR_=0.912, ρ_MA_=0.903) demonstrating replacement SNVs contribute greater noise to individual level risk precision in non-European ancestries.

We have demonstrated GRS2x provides a standardized and portable method to measure T1D polygenic risk across diverse datasets, addressing several critical limitations. Whilst future studies seek to improve discriminative performance, accessible and consistent measurement of T1D-PRS may provide greater immediate utility over marginal gains in performance alone. Though standardization is a crucial first step for clinical interpretation, validated reference ranges must be derived before the clinical utility of thresholds can be evaluated. Ancestry-specific refinement is also necessary: reduced performance in non-European ancestries may be partially driven by biases in EHR data or by ancestry-specific variation in allele frequencies, particularly HLA. Furthermore, precision of individual level risk estimates was observed to be more sensitive to SNV replacement in non-Europeans, underscoring the advantage of GRS2x as a unified approach eliminating the need for study-specific SNV modifications.

## Data Availability

GRS2x along with relevant diabetes polygenic scores are available through the Polygenic Risk Score Extension for Diabetes Mellitus (prsedm) package, distributed by GitHub (https://github.com/sethsh7/prsedm), PyPi (https://pypi.org/project/prsedm/) and Anaconda (https://anaconda.org/sethsh7/prsedm). Notebooks to deploy GRS2x on All of Us and UK Biobank are available on Github.

https://pypi.org/project/prsedm/

https://anaconda.org/sethsh7/prsedm

## Acknowledgements

A.M.L. is funded by a PhD studentship from Randox Laboratories Ltd. A.J.D. is funded by NIDDK K23DK140643. RAO is funded by NIDDK (R01DK121843, R01DK124395), Breakthrough T1D (4-SRA-2023-1375-M-B, 3-SRA-2022-1241-S-B, 2-SRA-2022-1261-S-B, 2- SRA-2022-1258-M-B, 2-SRA-2024-1620-S-B), The Leona M. and Harry B. Helmsley Charitable Trust (2016PG-T1D049, 2018PG-T1D049, 2103–05059, and G-2404-06858), and Randox Ltd. H.I.O. is funded by NIDDK T32DK007217 and Stanford Maternal and Childrens Health Research Institute. A.K.M. is supported by the FNIH with funding from AMP CMD RFP 2 and NHGRI U01HG011723. J.M.M. is supported by ADA grant #11-22-ICTSPM-16 and by NHGRI U01HG011723 and NIDDK R01DK137993 and U01 DK140757, AMP CMD award from RFP 6 from the FNIH, and a Medical University of Bialystok (MUB) grant from the Ministry of Science and Higher Education (Poland). M.A.R. is supported by NHGRI R01HG010140 and NIMH R01MH124244. M.S.U is funded by Doris Duke Clinical Scientist Development Award 2022063, NHGRI U01HG011723, and NIDDK (U54DK118612, UM1DK126185, U01DK140757). A.L.G. is funded by NIDDK (UM1-1DK126185, P30 DK116074). S.A.S. is funded by the Larry L. Hillblom Foundation (2024-D-017-FEL). We gratefully acknowledge All of Us participants for their contributions, without whom this research would not have been possible. We also thank the National Institutes of Health All of Us Research Program for making available the participant data examined in this study. This research has been conducted using the UK Biobank Resource under Application Number 9072.

## Author Contributions

A.M.L, R.A.O, M.N.W, A.L.G and S.A.S contributed to analysis and wrote the manuscript. R.A.O, A.L.G and S.A.S designed the study. A.J.D, H.I.O, D.P.F, K.A, A.K.M, J.M.M, M.A.R, M.S.U contributed to analysis and reviewed the manuscript. All authors contributed to the discussion and reviewed or edited the manuscript. S.A.S. is the guarantor of this work and, as such, had full access to all the data in the study and takes responsibility for the integrity of the data and the accuracy of the data analysis.

## Data and Resource Availability

GRS2x along with relevant diabetes polygenic scores are available through the Polygenic Risk Score Extension for Diabetes Mellitus (prsedm) package, distributed by GitHub (https://github.com/sethsh7/prsedm), PyPi (https://pypi.org/project/prsedm/) and Anaconda (https://anaconda.org/sethsh7/prsedm). Notebooks to deploy GRS2x on All of Us and UK Biobank are available on GitHub.

## References

1 Luckett, A. M. et al. Utility of genetic risk scores in type 1 diabetes. Diabetologia 66, 1589–1600 (2023). 10.1007/s00125-023-05955-y

2 Sims, E. K. et al. Screening for Type 1 Diabetes in the General Population: A Status Report and Perspective. Diabetes 71, 610–623 (2022). 10.2337/dbi20-0054

3 Sharp, S. A. et al. Development and Standardization of an Improved Type 1 Diabetes Genetic Risk Score for Use in Newborn Screening and Incident Diagnosis. Diabetes Care 42, 200–207 (2019). 10.2337/dc18-1785

4 Halldorsson, B. V. et al. The sequences of 150,119 genomes in the UK Biobank. Nature 607, 732–740 (2022). 10.1038/s41586-022-04965-x

5 Bick, A. G. et al. Genomic data in the All of Us Research Program. Nature 627, 340–346 (2024). 10.1038/s41586-023-06957-x

